# Validation of a guidelines-based digital tool to assess the need for germline cancer genetic testing

**DOI:** 10.1101/2024.05.07.24305624

**Authors:** Callan D. Russell, Ashley V. Daley, Durand R. Van Arnem, Andi V Hila, Kiley J. Johnson, Jill N. Davies, Hanah S. Cytron, Kaylene J. Ready, Cary M. Armstrong, Mark E. Sylvester, Colleen A. Caleshu

## Abstract

**Purpose:** Efficient and scalable solutions are needed to identify patients who qualify for germline cancer genetic testing. We evaluated the clinical validity of a brief, patient-administered hereditary cancer risk assessment digital tool programmed to assess if patients meet criteria for germline genetic testing, based on personal and family history, and in line with national guidelines.

**Methods:** We applied the tool to cases seen in a nationwide telehealth genetic counseling practice. Validity of the tool was evaluated by comparing the tool’s assessment to that of the genetic counselor who saw the patient. Patients’ histories were extracted from genetic counselor-collected pedigrees and input into the tool by the research team to model how a patient would complete the tool. We also validated the tool’s assessment of which specific aspects of the personal and family history met criteria for genetic testing.

**Results:** Of the 152 cases (80% ((121/152)) female, mean age 52.3), 56% (85/152) had a personal history of cancer and 66% (99/152) met genetic testing criteria. The tool and genetic counselor agreed in 96% (146/152) of cases. Most disagreements (4/6; 67%) occurred because the GC’s assessment relied on details the tool was not programmed to collect since patients typically don’t have access to the relevant information (pathology details, risk models). We also found complete agreement between the tool and research team on which specific aspects of the patient’s history met criteria for genetic testing.

**Conclusion:** We observed a high level of agreement with genetic counselor assessments, affirming the tool’s clinical validity in identifying individuals for hereditary cancer predisposition testing and its potential for increasing access to hereditary cancer risk assessment.

## INTRODUCTION

At least 5-10% of cancer is hereditary, arising from a germline pathogenic variant in a cancer predisposition gene^1^. 8% of individuals in the general population carry such pathogenic variants, yet the vast majority of these individuals do not know they possess this risk.^2–5^ Identification of individuals with a hereditary risk of cancer allows for personalized care such as more frequent and earlier cancer screening and risk-reducing surgeries. These measures have been shown to lead to earlier cancer diagnoses, improved prognosis, and/or prevention of cancer.^6–8^ In addition, among patients with cancer, identification of individuals with certain germline pathogenic variants is necessary for personalized cancer treatment such as PARP inhibitors for those with *BRCA1/2* mutations and immune checkpoint therapies for those with Lynch syndrome.^9, 10^ Given these clinical benefits, multiple professional guidelines recommend that oncologists, obstetricians-gynecologists and primary care doctors perform hereditary cancer risk assessment.^1, 6, 11–13^

However, there is ample evidence that in both non-specialty and oncology settings, hereditary cancer risk assessment is not performed as recommended by guidelines.^2, 4, 14–17^ Fewer than 20% of women with a history of breast or ovarian cancer who meet National Comprehensive Cancer Network (NCCN) criteria for germline cancer genetic testing have had such testing.^2^ Most individuals who haven’t had testing report never discussing testing with a healthcare provider.^2^ In a 2022 study of over 279,000 women receiving primary care at the Cleveland Clinic, only 22% of high-risk women had been referred for genetic testing.^4^ Additionally, that study found disparities in referrals based on race with Black individuals significantly less likely to be referred than White individuals. A 2023 study of over 1.3 million cancer patients found that while rates of germline genetic testing after a cancer diagnosis have increased over time, such testing remains heavily underutilized after a cancer diagnosis.^16^

Investigations into the reasons that providers do not perform guideline-recommended hereditary cancer risk assessments have revealed that providers perceive such assessments as valuable and important, but they face many barriers to performing them for their patients.^18–21^ Non-genetics providers feel they lack sufficient genetics expertise, do not feel confident answering patient questions related to genetic risk and genetic testing, and have difficulty staying up to date with advances in genetic testing ^18–21^. In addition, providers report they do not have time to adequately assess and counsel patients about hereditary cancer risk.^18, 21^ This is understandable, given it can take up to 30 minutes to collect the extensive family history that is often needed to determine if a patient meets guideline-based criteria.^22^ Furthermore, such criteria are complex and frequently change, making it difficult for providers to apply them. New approaches to hereditary cancer risk assessment are needed that address these barriers. Several paper-based screening tools have been developed; these are often brief forms completed by patients and scored by clinic staff. While they increase the identification of patients at high risk, they only cover a small subset of testing criteria and thus miss many patients who qualify for genetic testing.^23–26^

Digital tools have the potential to cover far more testing criteria and to assess patients in an automated fashion that does not depend on clinic staff. A variety of such solutions have arisen in recent years.^27–31^ This includes automated algorithms that leverage family history information already captured in the EHR^31^ as well as patient-facing digital tools that perform hereditary cancer risk assessment based on patient-entered personal and family history.^27, 28, 30^ Studies have found that such digital tools effectively identify at-risk patients who would have been overlooked.^25, 27, 30, 32–35^ Importantly, patients report high levels of satisfaction with digital tools in genomics care.^20, 27, 33, 36–38^ Thus, these digital tools show promise for helping increase access to guideline-recommended hereditary cancer risk assessment. However, appropriate validation of such tools must be performed to ensure the accuracy of their assessments.^20^ Of those that do report validation, there are mixed results on the sensitivity and specificity of tools. One such study showed 100% sensitivity and 99.5% specificity, however, the low-risk cases were fabricated and the validation only covered assessment for hereditary breast-ovarian cancer and Lynch syndrome^28^. Validation of another digital risk assessment tool found it failed to identify half of the individuals that genetics clinicians assessed to be at risk for a hereditary cancer predisposition^30^.

We developed RISE Risk Assessment Module: Hereditary Cancer to help providers perform hereditary cancer risk assessment without a significant time or process burden for them or their clinic staff (Figure 1). This is a brief, patient-administered web-based tool designed to assess whether germline cancer genetic testing may be indicated, consistent with national guidelines. The tool was created by a team of GCs with expertise in oncology, product managers, user experience designers, and software engineers. Patients answer questions about their personal and family history and the algorithm underlying the tool assesses whether that history indicates genetic testing is appropriate. To increase usability and efficiency, the history questions are programmed with skip logic so patients only see questions relevant to them. Unlike many existing hereditary cancer risk assessment tools, this tool was programmed to detect hereditary risk for a wide range of cancers, hereditary cancer syndromes, and tumors (Table 1). The algorithm is updated as guideline updates are released. The patient is immediately informed of the assessment result (Figure 1) and a separate PDF documenting the relevant history and assessment result is available to the provider in the platform. The platform is HIPAA-compliant and SOC2-certified.

**Figure 1.**
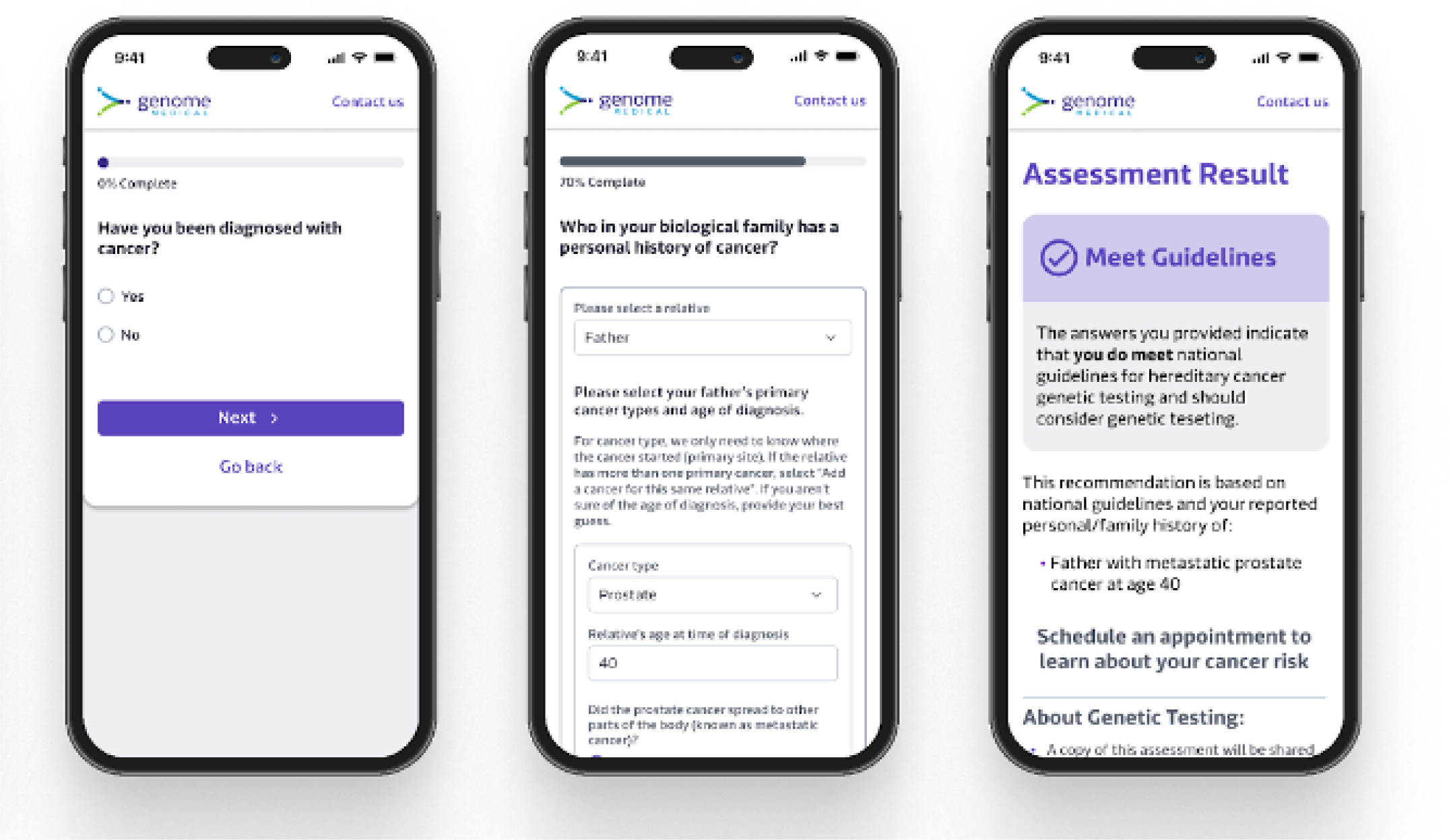
Screenshots of various steps in the digital tool including the welcome page, a history question, a family history question, and the assessment. Images courtesy of Genome Medical. Used with permission.

**Table 1:** RISE assesses hereditary risk for a variety of cancers, hereditary cancer syndromes, and tumors.

| **Cancers** | **Hereditary cancer syndromes** | **Tumors** |
| --- | --- | --- |
| Breast | Attenuated familial adenomatous polyposis (AFAP) | Paranganglioma |
| Colorectal | Classical familial adenomatous polyposis (FAP) | Pheochromocytoma |
| Gastric | Cowden |  |
| Kidney | Diffuse gastric cancer and lobular breast cancer |  |
| Medullary thyroid | Hereditary breast and ovarian cancer (HBOC) |  |
| Ovarian | Li-Fraumeni |  |
| Pancreatic | Lynch |  |
| Prostate | MUTYH-associated polyposis (MAP) |  |
| Uterine |  |  |

We sought to validate this hereditary cancer risk assessment tool against assessments made by genetic counselors (GCs) specialized in oncology, which is the current gold standard for hereditary cancer risk assessment.

## METHODS

To assess the clinical validity of the tool, the tool’s assessment of the patient meeting criteria for genetic testing was compared to the assessment made by the board-certified cancer GC who previously saw the patient for clinical care. The tool’s assessment was performed retrospectively and as part of this study only (not part of the patient’s clinical care).

### Patients

Cases were drawn from patients seen for pre-test genetic counseling for hereditary cancer risk in a nationwide telehealth genetic counseling practice between July 23, 2020, and October 23, 2020. We used purposive sampling to select cases that met criteria for genetic testing (as assessed by the GC who saw the patient for clinical care) to ensure that the sample covered the criteria most frequently invoked in clinical practice and for variance in cancer types. Cases that did not meet criteria for genetic testing (as assessed by the GC who saw the patient for clinical care) were consecutively selected.

### Data collection

Patient demographics, personal history of polyps or cancer, family history of cancer, and the GC’s assessment of whether the patient met criteria were extracted via retrospective review of the electronic medical record. The tool’s assessment was determined by the research team entering the patient’s personal and family history into the tool.

### Validation

Validity was operationalized as how often the tool’s assessment of whether the patient met criteria for genetic testing agreed with the assessment made by the GC who saw the patient clinically. When the tool’s assessment and the assessment made by the GC disagreed, a senior cancer GC (AD) reviewed the case to determine the origin of the disagreement.

We further assessed the performance of the tool in a subset of cases by determining the level of agreement between the tool and the research team on which aspects of a patient’s history met criteria.

The WIRB-Copernicus Group Institutional Review Board deemed the study exempt and approved a waiver of authorization for use and disclosure of protected health information (PHI) because the study analyzed deidentified secondary data.

## RESULTS

The dataset consisted of 152 patients seen for pre-test cancer genetic counseling with two-thirds meeting criteria for genetic testing (per GC assessment) and half having a personal history of cancer, with a variety of cancer types represented including breast, colorectal, uterine, prostate, ovarian, pancreatic, renal, melanoma, and skin (Table 2).

**Table 2:**
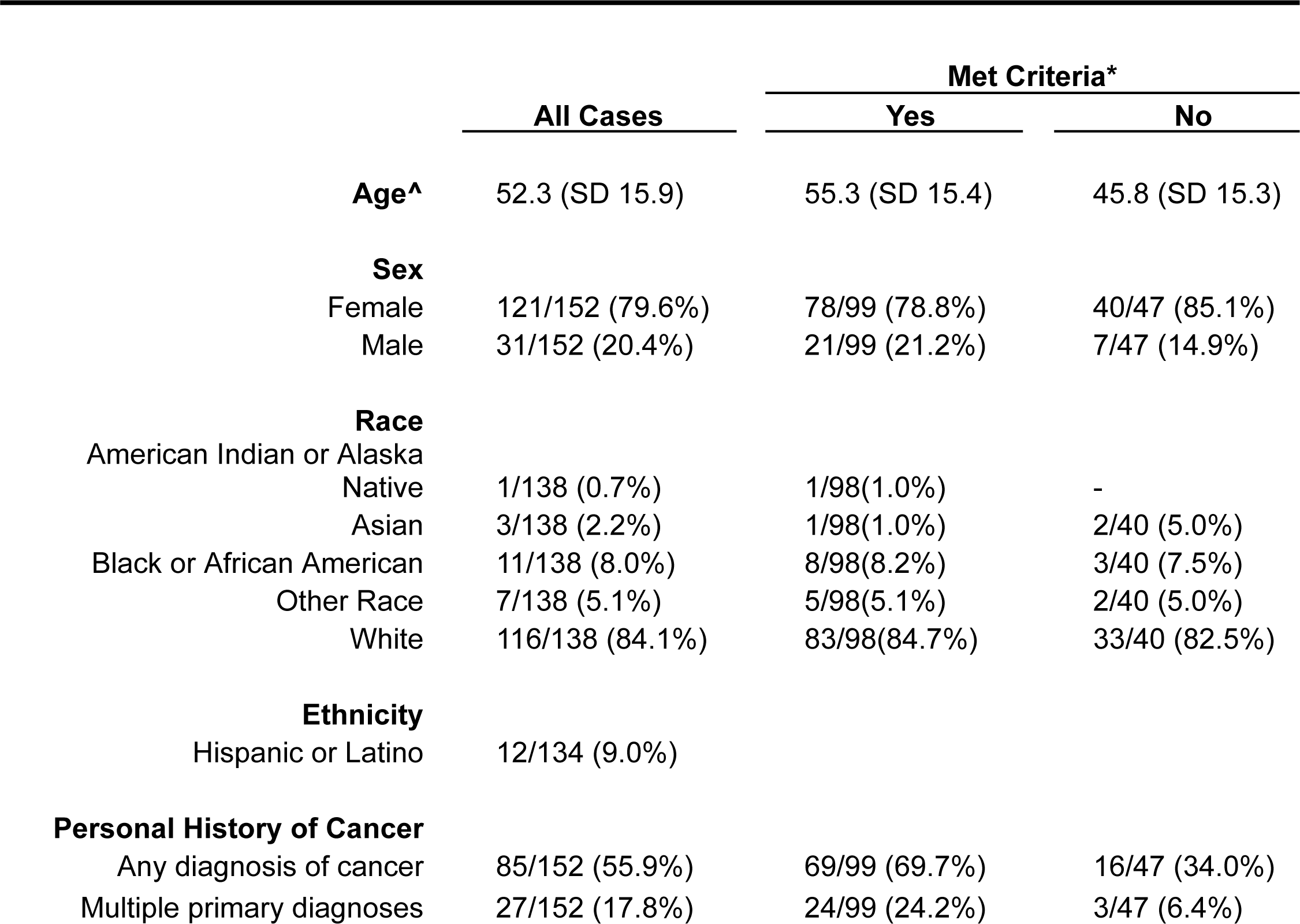
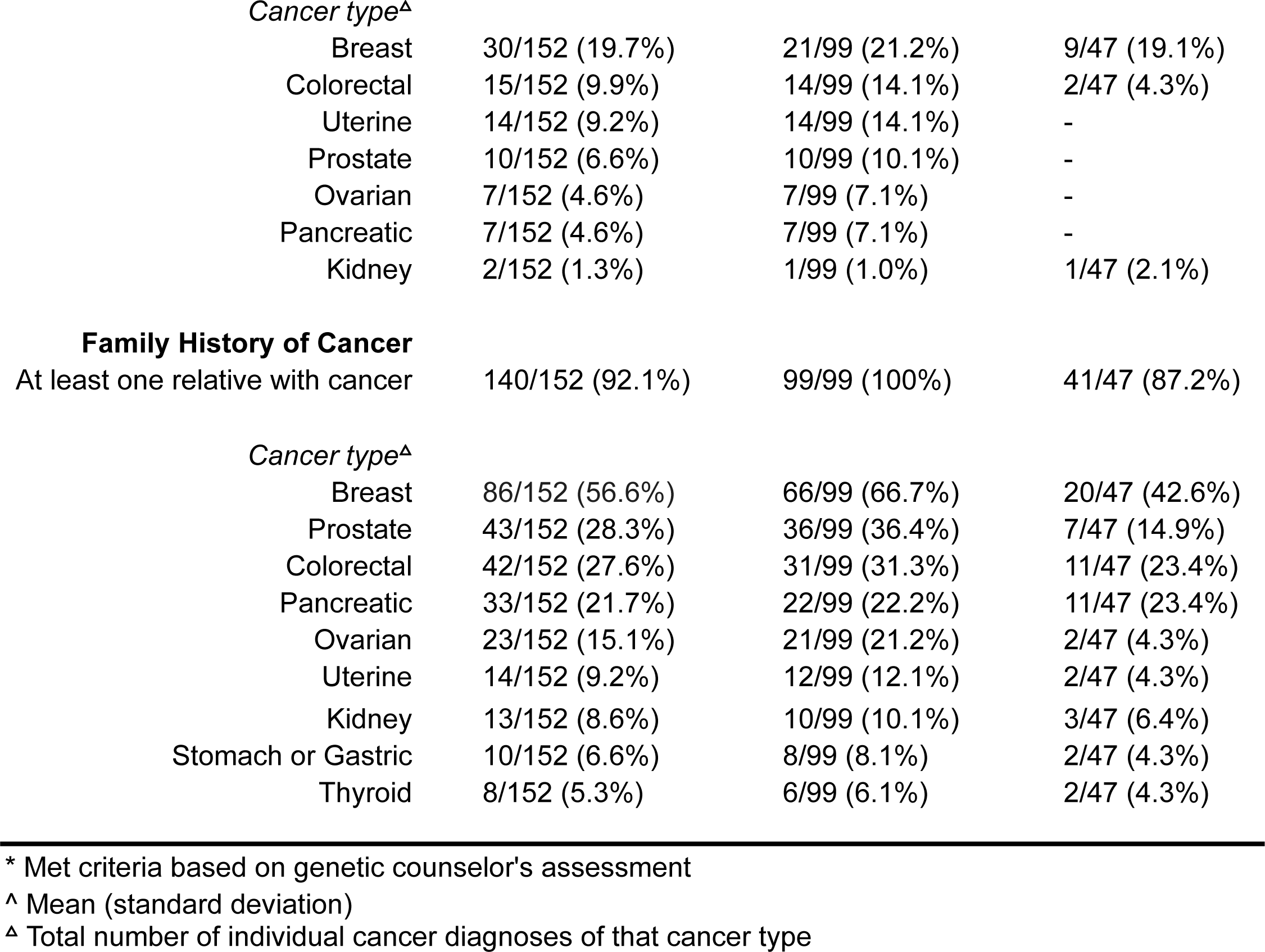
Patient Characteristics.

|  | All Cases | Met Criteria* |  |
| --- | --- | --- | --- |
|  |  | Yes | No |
| **Age^** | 52.3 (SD 15.9) | 55.3 (SD 15.4) | 45.8 (SD 15.3) |
| **Sex** |  |  |  |
| Female | 121/152 (79.6%) | 78/99 (78.8%) | 40/47 (85.1%) |
| Male | 31/152 (20.4%) | 21/99 (21.2%) | 7/47 (14.9%) |
| **Race** |  |  |  |
| American Indian or Alaska Native | 1/138 (0.7%) | 1/98(1.0%) | - |
| Asian | 3/138 (2.2%) | 1/98(1.0%) | 2/40 (5.0%) |
| Black or African American | 11/138 (8.0%) | 8/98(8.2%) | 3/40 (7.5%) |
| Other Race | 7/138 (5.1%) | 5/98(5.1%) | 2/40 (5.0%) |
| White | 116/138 (84.1%) | 83/98(84.7%) | 33/40 (82.5%) |
| **Ethnicity** |  |  |  |
| Hispanic or Latino | 12/134 (9.0%) |  |  |
| **Personal History of Cancer** |  |  |  |
| Any diagnosis of cancer | 85/152 (55.9%) | 69/99 (69.7%) | 16/47 (34.0%) |
| Multiple primary diagnoses | 27/152 (17.8%) | 24/99 (24.2%) | 3/47 (6.4%) |
| *Cancer type<sup>Δ</sup>* |  |  |  |
| Breast | 30/152 (19.7%) | 21/99 (21.2%) | 9/47 (19.1%) |
| Colorectal | 15/152 (9.9%) | 14/99 (14.1%) | 2/47 (4.3%) |
| Uterine | 14/152 (9.2%) | 14/99 (14.1%) | - |
| Prostate | 10/152 (6.6%) | 10/99 (10.1%) | - |
| Ovarian | 7/152 (4.6%) | 7/99 (7.1%) | - |
| Pancreatic | 7/152 (4.6%) | 7/99 (7.1%) | - |
| Kidney | 2/152 (1.3%) | 1/99 (1.0%) | 1/47 (2.1%) |
| **Family History of Cancer** |  |  |  |
| At least one relative with cancer | 140/152 (92.1%) | 99/99 (100%) | 41/47 (87.2%) |
| *Cancer type<sup>Δ</sup>* |  |  |  |
| Breast | 86/152 (56.6%) | 66/99 (66.7%) | 20/47 (42.6%) |
| Prostate | 43/152 (28.3%) | 36/99 (36.4%) | 7/47 (14.9%) |
| Colorectal | 42/152 (27.6%) | 31/99 (31.3%) | 11/47 (23.4%) |
| Pancreatic | 33/152 (21.7%) | 22/99 (22.2%) | 11/47 (23.4%) |
| Ovarian | 23/152 (15.1%) | 21/99 (21.2%) | 2/47 (4.3%) |
| Uterine | 14/152 (9.2%) | 12/99 (12.1%) | 2/47 (4.3%) |
| Kidney | 13/152 (8.6%) | 10/99 (10.1%) | 3/47 (6.4%) |
| Stomach or Gastric | 10/152 (6.6%) | 8/99 (8.1%) | 2/47 (4.3%) |
| Thyroid | 8/152 (5.3%) | 6/99 (6.1%) | 2/47 (4.3%) |

\* Met criteria based on genetic counselor's assessment
<sup>Δ</sup> Mean (standard deviation)
<sup>Δ</sup> Total number of individual cancer diagnoses of that cancer type

In 96% (146/152) of cases, the tool’s assessment of whether the patient met criteria for genetic testing agreed with the GC’s assessment (Figure 2). Among patients who met criteria (by GC assessment), there was 95% (94/99) agreement between the tool and GC. Among patients who did not meet criteria (by GC assessment), there was 98% (46/47) agreement between tool and GC.

**Figure 2.**
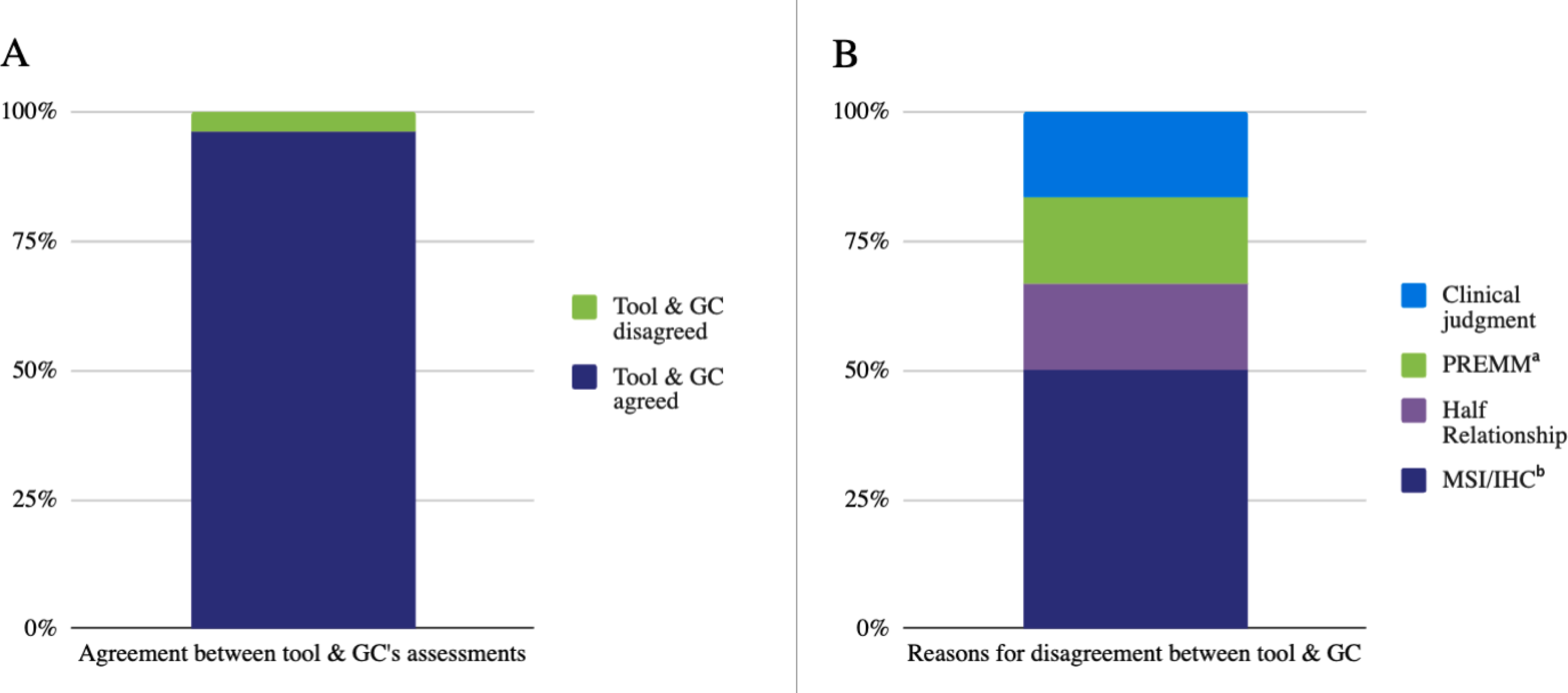
High level of agreement between tool and genetic counselor (A) Stacked bar chart showing the percentage of cases with agreement in assessments made by the tool and the GC (96% (146/152)) (B) Reasons for disagreement in the 3.9% (6/152) of cases were the tool’s and GC’s assessments differed. The tool does not ask about clinical details patients typically cannot report such as MSI/IHC and risk of having a germline pathogenic variant based on the PREdiction Model for gene Mutations (PREMM5), nor does it account for half relationships. In one case the GC’s application of their clinical judgment in interpreting a patient’s history of polyps contributed to disagreement. a. PREMM = PREdiction Model for gene Mutations. b. MSI/IHC = Microsatellite Instability/Immunohistochemistry.

For the cases (3.9% (6/152)) where there was disagreement between the tool and GC, we examined why the assessments differed. Most differences in assessment (67% (4/6)) occurred because the GC assessment depended on a specific aspect of history that the tool did not ask about. This included microsatellite instability/immunohistochemistry (MSI/IHC) results (50% (3/6)) and risk of having a germline pathogenic variant based on the PREdiction Model for gene Mutations (PREMM5; 16.7% (1/6))^39^. Disagreement occurred in one case because the GC applied their clinical expertise in interpreting the patient’s polyp history as meeting criteria based on likely polyp type, while the tool assessed the patient as not meeting criteria because polyp type was unknown (16.7% (1/6)). The final case of disagreement arose because the tool does not currently allow entry of half-relationships (16.7% (1/6)).

Performance of the tool depends on the accuracy of the underlying algorithm in assessing that specific aspects of the patient’s personal and/or family history meet criteria. To further validate the tool, we compared the tool’s and research team’s assessment of which specific aspects of the patient’s history met criteria for a subset of cases (40.8% (62/152)). This involved comparing the history-based rule(s) in the tool’s algorithm that were triggered for each case to the research team’s assessment of which history-based rule(s) should have been triggered. This subset of cases had similar characteristics to the overall sample (Supplemental Table 1), with the exception that they all met criteria for genetic testing (as assessed by the GC). In all cases, the research team and the tool agreed on (100% (62/62)) which aspects of the patient’s personal and family history met criteria for genetic testing. Across these 62 cases, specific aspects of the patient’s history were recognized by 65 different rules in the tool’s algorithm, and these rules were triggered a total of 269 times across the 62 patient cases, with complete agreement between the tool and research team each time they were triggered (100% (269/269)). Each rule was triggered by a mean of 3.8 cases (SD 3.5) and the mean number of rules triggered per case was 4.1 (SD 2.7). All algorithm rules that are used in clinical practice with the highest frequency were validated individually (100% (37/37)), as were most intermediate frequency rules (68.4% (26/38)) (Table 3). The majority of individual rules in the following cancer types were validated: breast, ovarian, pancreatic (71.7% (33/46)); colorectal, endometrial (93.3% (14/15)); prostate (69.6% (16/23)) (Table 3).

**Table 3:**
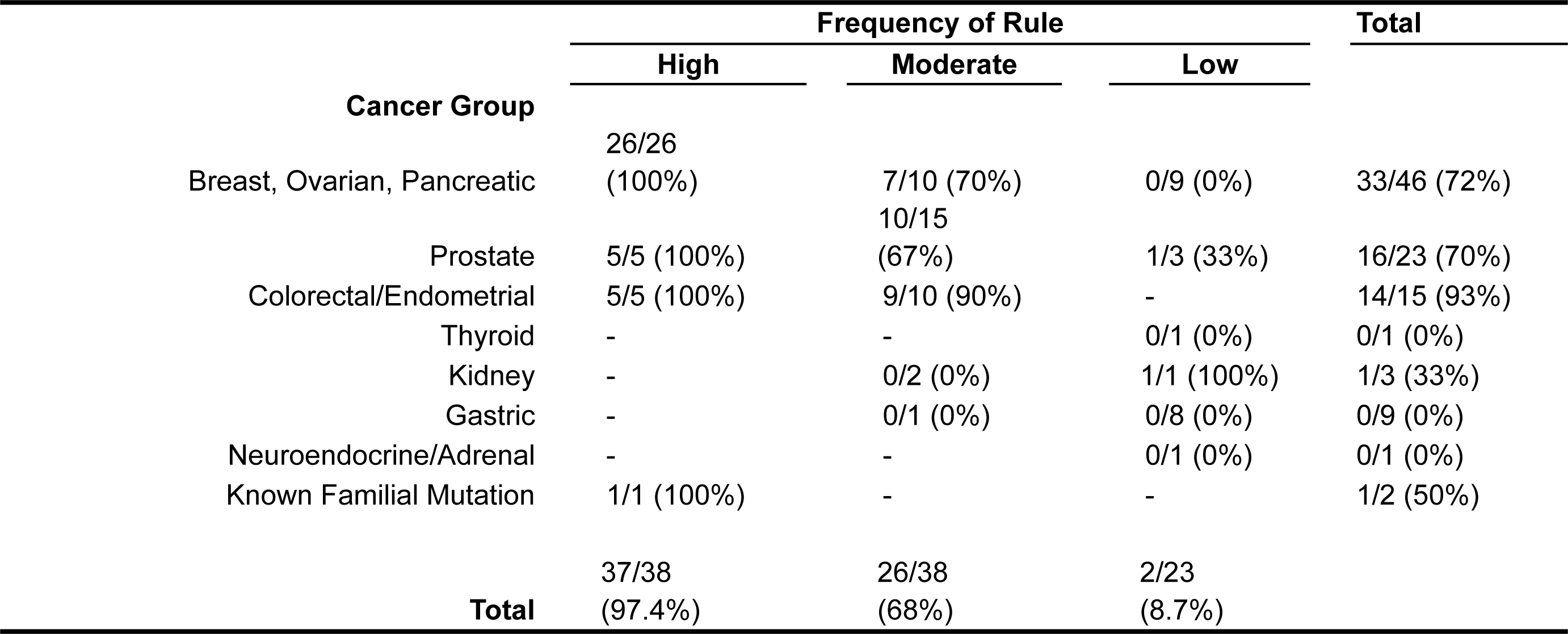
Rules Validated.

## DISCUSSION

We observed a high level of agreement between the tool and GCs, which suggests that the tool is accurate in its assessments of whether patients meet criteria for genetic testing. The rate of agreement we observed was markedly higher than that seen by Cohn et al, similar to that seen by Baumgart et al and slightly lower than that reported by Bucheit et al^28, 30^. It is also at the high end of the range of accuracy reported by USPTF in their review of several less automated hereditary cancer risk assessment tools^40^. Furthermore, we found complete agreement between the tool and the study team on which specific aspects of a patient’s personal and family history meet criteria. This is particularly critical when hereditary cancer risk assessment is done in primary care or other population-based settings since many unaffected patients in such settings qualify for genetic testing based on just one aspect of their family history^35^. Taken together, these findings suggest the tool has an acceptable level of accuracy that is higher than or comparable to other risk assessment tools. It is also notable that the tool was validated using cases meeting criteria for a variety of hereditary cancer predispositions. Other digital risk assessment tools, and also validation of those tools, have primarily focused only on BRCA1/2 and Lynch syndrome ^27, 28, 30^. In contrast, the current validation covered risk for a wide range of cancers, cancer syndromes, and tumors, all of which the tool is programmed to detect (Table 1). Another strength of this study is the use of real patient cases, in contrast to prior work on validation of digital risk assessment tools which has relied, at least in part, on fictitious cases^28^.

It is worth considering the minority of cases where the tool and GC disagreed. Of note, none of the disagreements were due to errors in the functioning of the tool. Most disagreements occurred because of history questions that were intentionally left out of the tool (ex. MSI/IHC (3 cases), PREMM5 (1 case)) due to our clinical experience that patients do not have the necessary information to answer such questions. Asking more questions and questions patients can’t answer can increase cognitive burden and decrease usability, both of which have been shown to decrease patient engagement with digital health tools^41,42^. RISE was intentionally designed to be brief to maximize completion rates; we’ve found that more than 95% of patients complete the tool, with most patients completing the tool in less than 3 minutes^35^. Given that MSI/IHC contributed to disagreement in multiple cases, we could add a question on that to the tool and then study whether patients can answer it and whether completion rates decrease. An additional area for improvement of the tool is the addition of half-relationships, as this contributed to disagreement in one case.

The high level of agreement between the tool and GCs that we observed, combined with prior research on feasibility and acceptability of genomics digital tools,^27, 33, 36–38, 43–45^ supports them as promising solutions to increasing access to hereditary cancer risk assessment without burdening clinicians. In a recent systematic review, Lee et al found that 84% of 87 studies on digital tools in genomics reported a positive outcome and that digital tools increased provider efficiency and decreased the time providers need to spend with patients ^36^. Hereditary cancer risk assessment tools could also make periodic re-assessment more feasible, which is recommended by guidelines.^6^ While recent studies demonstrate that such tools effectively identify at-risk patients who were otherwise un-ascertained, they also find that additional implementation work is needed to increase the proportion of these at-risk patients who go on to have genetic testing.^46, 47^ This demonstrates that innovation and practice improvement are needed at multiple steps in the care pathway to ensure access to the benefits of genomic medicine.

An important limitation of this work is that patient histories were not entered by patients themselves, but instead by the research team. While the validation covered a range of aspects of history and types of hereditary cancer risk, it was not exhaustive; we did not validate every rule in the algorithm or every way a given criteria could be met. Additionally, since the cases included in our study had already been assessed as needing genetic counseling, they are not representative of a lower-risk population. We did not have sufficient variance in disagreement in assessment or either race or ethnicity to be able to investigate disparities in the tool’s performance. Multiple studies have found disparities in cancer genetics care based on race^48, 49^, ethnicity, and socioeconomic status ^4^. While digital tools have potential to reduce disparities, care in their design, implementation, and evaluation is needed to ensure they perform and benefit patients equitably. Finally, our sampling method did not allow for calculation of sensitivity and specificity.

## CONCLUSION

We observed a high degree of agreement between the digital tool and cancer genetic counselors’ assessments of whether patients meet criteria for germline genetic testing. Combined with prior findings on feasibility, acceptability, and efficiency of digital tools in genomics, our results suggest that RISE Risk Assessment Module: Hereditary Cancer could help increase access to hereditary cancer risk assessment and genetic testing without significantly burdening clinicians.

## Data Availability

All data produced in the present study are available upon reasonable request to the authors

## ACKNOWLEDGEMENTS

We thank Caitlin Campbell for assistance with writing and both Isabela Dall’Oglio Bucco and Cecilia Kessler for administrative assistance.

## SUPPLEMENTAL MATERIAL

**Supplemental Table 1:**
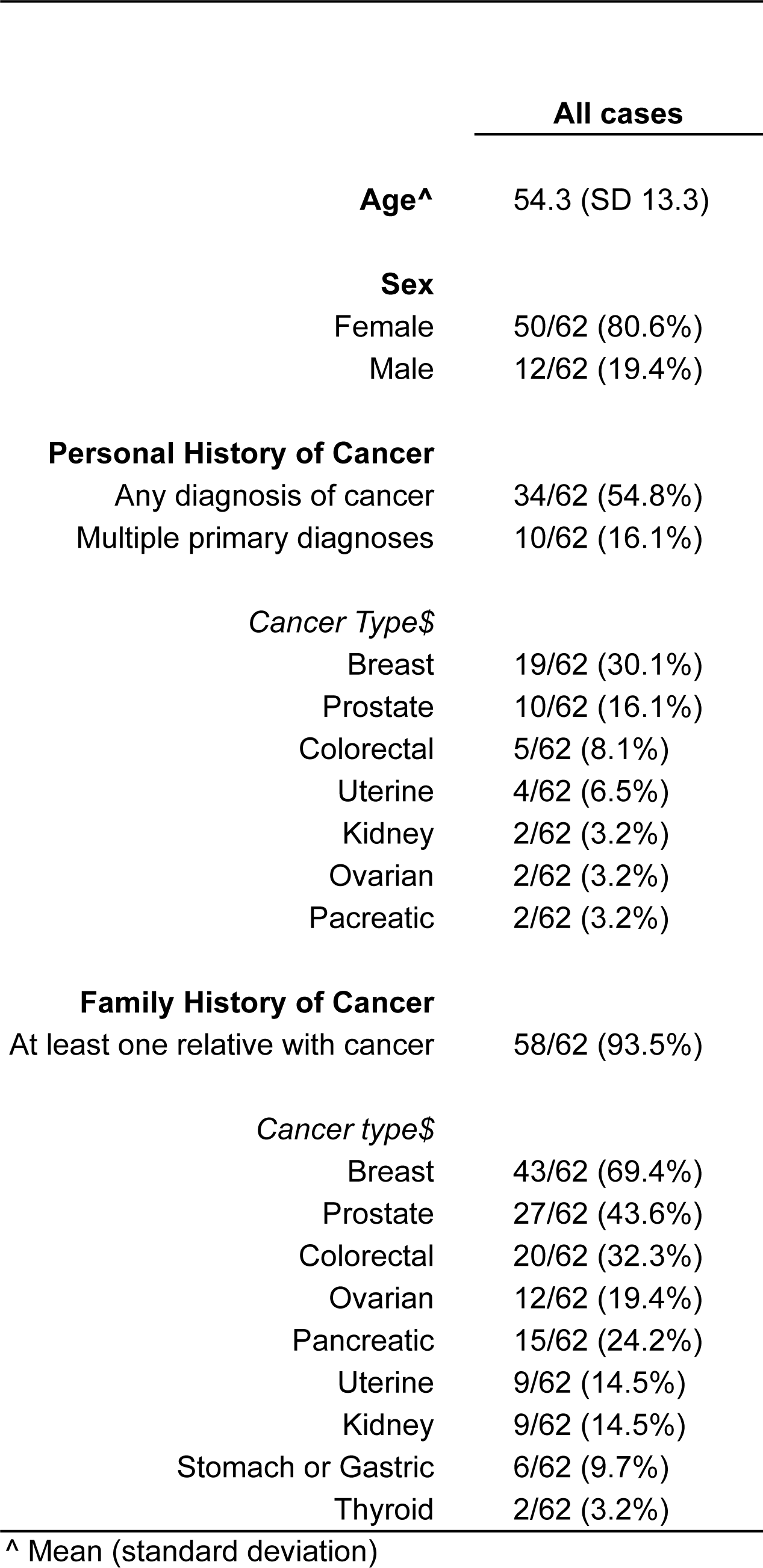
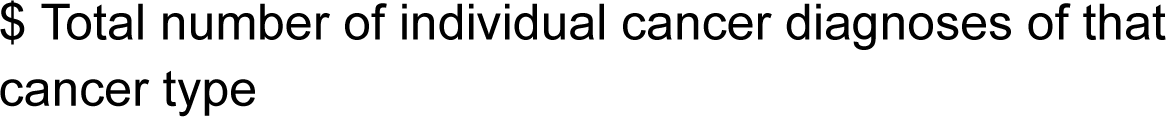
Characteristics of Cases Used for Rule Validation.

|  | All cases |
| --- | --- |
| **Age<sup>^</sup>** | 54.3 (SD 13.3) |
| **Sex** |  |
| Female | 50/62 (80.6%) |
| Male | 12/62 (19.4%) |
| **Personal History of Cancer** |  |
| Any diagnosis of cancer | 34/62 (54.8%) |
| Multiple primary diagnoses | 10/62 (16.1%) |
| *Cancer Type\$* | |
| Breast | 19/62 (30.1%) |
| Prostate | 10/62 (16.1%) |
| Colorectal | 5/62 (8.1%) |
| Uterine | 4/62 (6.5%) |
| Kidney | 2/62 (3.2%) |
| Ovarian | 2/62 (3.2%) |
| Pancreatic | 2/62 (3.2%) |
| **Family History of Cancer** |  |
| At least one relative with cancer | 58/62 (93.5%) |
| *Cancer type\$* | |
| Breast | 43/62 (69.4%) |
| Prostate | 27/62 (43.6%) |
| Colorectal | 20/62 (32.3%) |
| Ovarian | 12/62 (19.4%) |
| Pancreatic | 15/62 (24.2%) |
| Uterine | 9/62 (14.5%) |
| Kidney | 9/62 (14.5%) |
| Stomach or Gastric | 6/62 (9.7%) |
| Thyroid | 2/62 (3.2%) |

<sup>^</sup> Mean (standard deviation)
\$ Total number of individual cancer diagnoses of that cancer type

